# Designing for Success: A Prospective Evaluation of Implementation Factors Affecting a Prototype Novel Medical Device in a Low-Resource Environment Using the CFIR 2.0

**DOI:** 10.64898/2026.02.03.26345034

**Authors:** Megan Morecroft, Ulziikhishig Byambabayar, Natalie DeMello, Annamarie Saarinen, Emma Rees

## Abstract

1

**Background:** Implementation challenges are a major contributor to the failure of novel medical technologies in low-resource settings. Although frameworks such as the Consolidated Framework for Implementation Research (CFIR) are widely used to evaluate interventions post-implementation, their prospective application during the product development phase remains limited.

**Methods:** This study aimed to prospectively assess implementation factors relevant to the future adoption of a prototype handheld ultrasound device designed to improve congenital heart disease (CHD) screening in low- and middle-income countries (LMICs). Structured observational field notes were collected during a clinical site visit to Mongolia in May 2024. Observations were made across five hospitals and two high-level stakeholder meetings. Data were thematically analysed using Braun and Clarke’s reflexive method, followed by deductive mapping to CFIR 2.0 domains.

**Results:** The final codebook included 70 codes mapped to four CFIR 2.0 domains: Innovation, Individuals, Inner Setting, and Outer Setting. Key facilitators included high staff motivation, alignment of hospital and national goals with device priorities, and the device’s simplified screening procedure and cloud-based data transfer capability. Identified barriers included inconsistent infrastructure across hospitals, limited access to trained staff, lack of maintenance systems, and fragmented digital health records.

**Conclusions:** Prospective application of CFIR 2.0 provided a systematic method for identifying factors likely to influence implementation success. This study demonstrates the value of incorporating implementation science early in device development, particularly in LMIC contexts where design-stage decisions can determine device success. By embedding structured implementation analysis into product development, developers may improve alignment between design and setting, reduce downstream failures, and accelerate adoption of new technologies.

## 2 Introduction

Congenital heart disease (CHD) is the most common congenital defect worldwide, affecting approximately 1 in every 100 births. It is a leading contributor to newborn mortality and is the primary cause of congenital abnormality-related deaths [1]. In 2021, the global mortality for CHD was over 250,000, with 167,000 of these deaths occurring in infants under 1 year of age [2]. Early detection, therefore, is crucial to improving outcomes; in the case of critical CHD, early diagnosis can reduce mortality from 27% to 16% [1, 3].

The first opportunity for CHD diagnosis arises during prenatal ultrasound screening for fetal anomalies, often referred to as the “20-week anomaly scan”. Whilst developments in ultrasound technology and prenatal screening programmes have improved rates of prenatal CHD diagnosis worldwide, detection rates remain highly variable between countries and regions, especially in low and middle-income countries (LMICs) [4-6].

A survey of 32 centres across 11 LMIC countries within the International Quality Improvement Collaborative for CHD found that only 19% of centres consistently included CHD screening as part of routine antenatal care [5]. Given that approximately 45% of CHD cases in the UK are not detected prenatally despite an extensive prenatal screening programme, the number of infants born undiagnosed in lower-resource counties is likely to be large [7-10].

For those infants born undiagnosed, many countries rely on newborn screening programmes to identify CHD [11]. The adoption of pulse oximetry, a low-cost and low-resource method for screening, is increasingly enabling the detection of CHD after birth .in countries where it is available [12-15]. However, screen-failure infants still require a full ultrasound of the heart (echocardiogram) to determine a CHD diagnosis.

In LMICs, this may represent a significant challenge; equipment and appropriately trained medical staff can be absent or large distances away from rural communities, with poor access to transportation [5, 16]. Many LMIC centres that do have access to imaging equipment additional barriers related to lack of portable machines, age-appropriate equipment (such as small ultrasound probes), and appropriately trained staff [5].

The culmination of these barriers contributes heavily to the mortality burden of CHD in LMICs; in 2016, an estimated 165,000 LMIC children with CHD died a preventable death, totalling 96.4% of excess CHD-related child mortality worldwide [17].

To address this diagnostic gap, Bloom Standard Inc. is developing a prototype handheld ultrasound device designed to bridge pulse oximetry screening and difficult-to-access full echocardiography. The goal is to identify infants who require urgent referral while avoiding unnecessary transfers for those with normal or low-risk findings, by enabling frontline healthcare workers to perform simplified cardiac scans with minimal training.

While technological innovations like this hold promise, their success depends not only on clinical performance but also on successful integration into the healthcare system. Although usability and human factors are often considered in device design and regulatory approval, there is little evidence that manufacturers prospectively plan for broader implementation challenges such as cost, supply chains, workforce capacity, or integration into existing health systems [18-21].

This disconnect can have serious consequences. It is estimated that 40–70% of medical devices in low-resource settings are broken, unused, or unfit for purpose [22]. Given that the average development cost of a medical device cleared via the FDA 510(k) pathway exceeds $30 million, addressing implementation barriers early is not only a health system imperative but also an economic one [23].

Implementation science offers a way to anticipate these barriers and design for success by understanding how innovations such as medical devices can be effectively adopted and used within routine practice [24]. Among the most widely used tools in this field is the Consolidated Framework for Implementation Research (CFIR), recently updated to CFIR 2.0. The framework offers a structured, multi-level approach to evaluating the complex interactions between the intervention (here, a medical device), individuals, organisations, and wider environmental and social context [19, 25-28].

In this study, we apply CFIR 2.0 prospectively to examine the potential implementation of Bloom Standard’s prototype device in a LMIC context. To our knowledge, this is the first published use of CFIR 2.0 to evaluate a device still in the development/pre-market phase. By integrating implementation insights into the early design process, we aim to inform both product design and strategies for sustainable implementation.

## 3 Methods

### 3.1 Aims

This study aimed to prospectively identify and evaluate implementation factors likely to influence the adoption of a novel medical device in a LMIC setting. By undertaking implementation analysis early in the product development process, the goal was to inform future device design and maximise deployment efficiency.

### 3.2 Design

This research employed a qualitative, observational design using structured ethnographic field notes collected during an observational field visit to Mongolia. The prototype device was being used in clinical settings as part of a separate ethically approved clinical investigation. Field notes were taken across five hospital sites and during two official stakeholder meetings.

For full details on the context of the trip and the template field note form see Supplementary File 1.

#### Participants and Setting

All data were collected by a member of the research team during a site visit to Mongolia in May 2024. Observations occurred in public, non-clinical and clinical areas with full awareness of local staff and stakeholders. No formal interviews or interactions with healthcare staff or patients were conducted, and no identifiable personal data were collected. This study was restricted to passive observation by researchers invited to attend clinical and administrative settings.

#### Data Collection and Sources

Data were gathered using a structured field note template that included prompts on setting characteristics, summary of key conversations or presentations, and device use in practice. During each hospital visit or meeting, the observer recorded detailed descriptive notes. After each session, notes were summarised and accompanied by a reflexive commentary.

This approach allowed capture of contextual insights, staff reactions, and implementation-relevant observations across multiple settings.

#### Data Analysis

Analysis followed Braun and Clarke’s six-phase approach to reflexive thematic analysis [29-31], chosen for its flexibility and alignment with interpretive qualitative methodologies. This method is particularly suited for contexts where the researcher’s positionality may influence data interpretation and has been used by others in the analysis of qualitative data for medical device design [21, 32, 33].

Field notes were transcribed to digital form and imported to NVIVO 10 software, which was used to iteratively code each set of notes and develop themes. Codes were regularly reviewed for relevance and overlap, with potential themes noted in the research diary. Once the full dataset had been analysed the codes were reviewed and grouped into themes and sub-themes.

When the first round of theming was complete, the results were deductively re-reviewed within the context of the CFIR 2.0. Codes and themes were subsequently re-grouped into relevant CFIR domains and sub-domains.

Final codes and themes were reviewed in the context of the research aims to ensure the output was meaningful in addressing the research questions. Codes deemed not relevant to the research aims were sorted under a “Not Relevant” heading within the software for transparency purposes. Performing the initial coding and theming independent of the framework domains reduced the analysis bias, whilst the subsequent deductive re-grouping of the data helped frame the analysis in line with the study aims and research question.

#### Participants and Ethical Considerations

No ethical approval was sought for this research as data collection did not require the participation of any individuals outside of the research team, and all field notes were collected in a non-invasive and non-interactive manner in a public setting. Infants involved in device testing were part of a separate, ethically approved research study^1^ run by Bloom Standard Inc. and their clinical collaborators in Mongolia. Their participation was subject to informed consent by their parent(s) or guardian(s) and included the taking of pictures and videos by members of the Bloom Standard, Swansea University, and Mongolia clinical team for commercial and research use.

While formal ethical approval was not required due to the non-interventional nature of this study, institutional leaders at each site were informed of the purpose of data collection, and permission was obtained to observe clinical and administrative activities. Observations were conducted openly, and field notes did not include any personal identifiers or private clinical data. The research team acknowledges the potential for power imbalances in international health partnerships and aimed to minimise disruption and maintain respect for local workflows. The study was conducted in collaboration with Mongolian clinical partners, who contributed to site access, contextual interpretation, and manuscript preparation. Findings will be shared with local stakeholders to inform device implementation planning and further research.

#### Reporting Guidelines

This study is reported in accordance with the Standards for Reporting Implementation Studies (StaRI) guidelines [34]. Given the qualitative and observational design, relevant elements of the COREQ checklist were also considered. A completed StaRI checklist is provided as Supplementary Material.

## 4 Results

The final codebook comprised 70 codes and sub-codes mapped to four of the five CFIR 2.0 domains: the Innovation, Individuals, Inner Setting, and Outer Setting. The Process domain was not applicable due to the pre-implementation nature of the study.

**Table 1.** Summary of CFIR 2.0 Domains and Sub-Domains Identified.

| Domain | Sub-domain |
| --- | --- |
| **The Innovation** | Innovation Design |
|  | Innovation Relative Advantage |
| **Individuals** | Motivation |
| **Inner Setting** | Communications |
|  | Mission Alignment |
|  | Structural Characteristics |
|  | Tension for Change |
| **Outer Setting** | Local Conditions |

**Figure 1.**
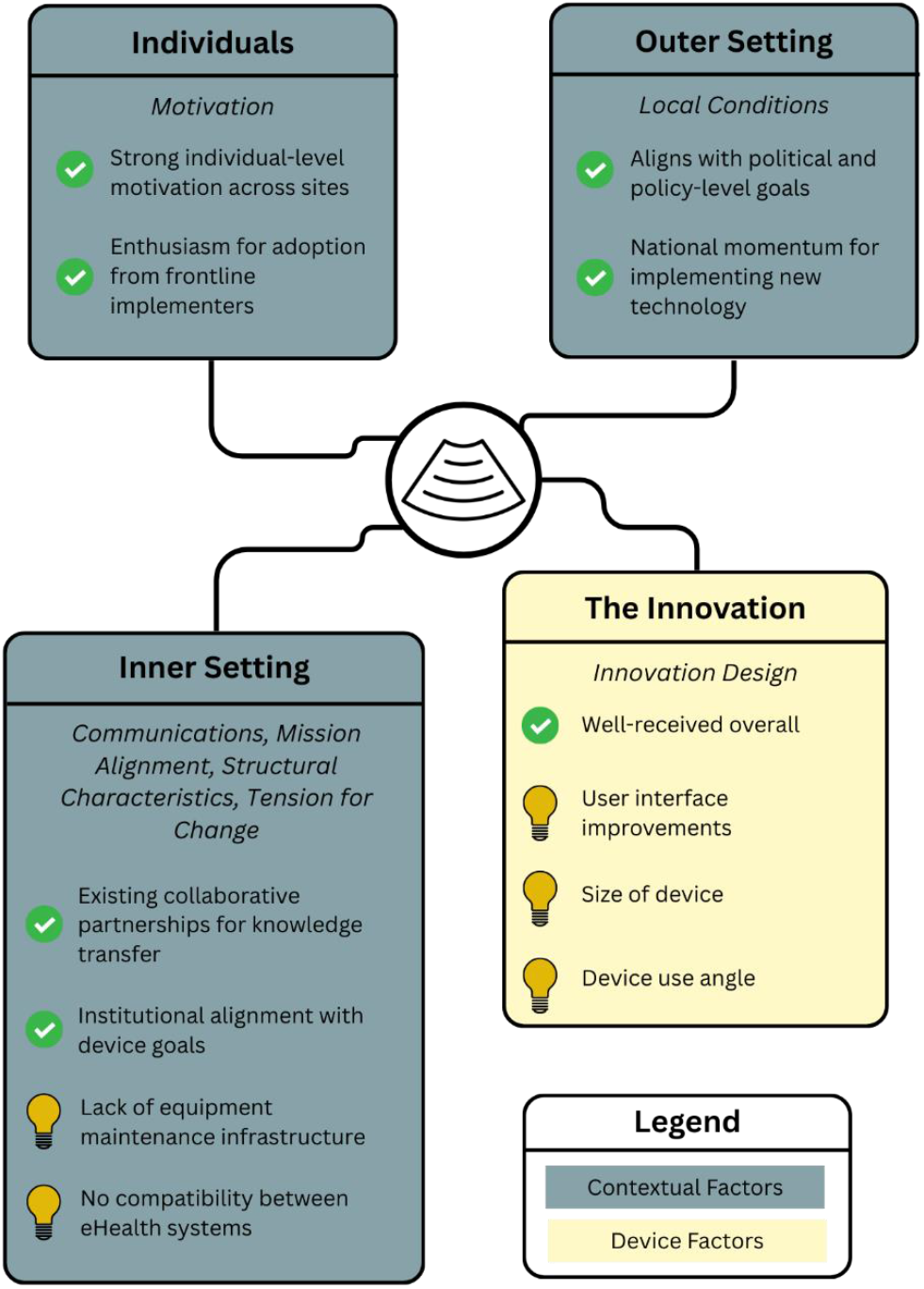
Visual summary diagram of results mapped to CFIR 2.0 domains

### 4.1 Individuals

#### Sub-domain: Motivation

Across sites, strong individual-level motivation was evident among clinicians and staff. Observations consistently recorded high levels of interest in the device and its potential utility:

> *“The scanning is done in a very small room attached to the meeting space. There was limited space to observe so the room quickly became crowded by staff who wanted to see the device working.” – Hospital 4*
>
> *“The [Neonatal Intensive Care Unit] staff were excited by the device and were very keen to see it being used on the baby” – Hospital 5*

This enthusiasm was evident regardless of setting and suggests early eagerness for adoption from frontline implementers.

### 4.2 Inner Setting

The hospital context, representing the primary setting for implementation, generated themes across four CFIR sub-domains.

#### Sub-domain: Communications

Collaborative partnerships were active in several hospitals, particularly focused on staff training and development:

> *“The hospital has large collaboration projects with surrounding countries to offer training abroad and bring foreign doctors in to train local doctors.” – Hospital 4*
>
> *“The hospital is collaborating with the [World Health Organisation] to reduce their under 5yo mortality and morbidity rate by 2030*… *ongoing projects with the WHO including offering specialised training to clinicians from the countryside*… *exchange programme with a Russian hospital*…*” – Hospital 5*

Such partnerships may facilitate knowledge transfer and support implementation efforts.

#### Sub-Domain: Mission Alignment

Hospitals demonstrated mission alignment with the device’s objectives, especially relating to early detection and health outcome improvement:

> *“The hospital has been making big efforts to improve their service in recent years. Since 2020, they have had 22 new ambulances and have decreased the <5yo mortality rate by 50% compared to 2016.” – Hospital 4*
>
> *“The hospital is part of the pulse oximetry screening programme, having implemented pulse ox within 5 months of the Newborn Foundation’s first visit.” – Hospital 2*

These observations suggest institutional readiness and alignment with innovation goals.

#### Sub-Domain: Structural Characteristics

Significant differences in digital infrastructure and physical resources were observed between hospitals:

- eHealth systems ranged from fully paper-based to partially digitised:

> *“Hospital 1 was recording all patient interactions by hand in a book*…*”*
>
> *“The hospital moved to eHealth in Aug 2021. Currently, half of records are digital and half remain paper. They aim to be completely digital within 2 years.” – Hospital 3*

- Physical infrastructure varied widely. Some facilities had minimal functioning equipment, whereas others were better resourced:

> *“Staff in hospitals 1 and 2 only had access to a single new ultrasound machine, whereas staff in hospital 3 had access to 4 or 5 ultrasound machines and 4 echocardiography machines.”*
>
> *“Compared to the hospitals visited on day 1 and 2, [hospital 2] has improved facilities and equipment*… *However, the hospital is in poor repair in many of the areas that we saw (broken flooring and ceiling tiles, extremely limited toilet and handwashing facilities that often did not have soap or toilet paper)*… *Staff say repairs are difficult to undertake as funds are so limited*… *equipment typically comes from donations*…*” – Hospital 2*
>
> A major concern among many of the staff members was the lack of maintenance infrastructure:
>
> *“Manufacturers of medical equipment do not train and employ maintenance employees within the country, eliminating any possibility of maintenance contracts.” – Hospital 3*

#### Sub-Domain: Tension For Change

Despite these infrastructure limitations, there was widespread recognition of unmet need and a strong drive for change. Hospitals faced significant challenges in providing follow-up echocardiography for screen-failed infants due to equipment shortages and staff training gaps:

> *“Have one old echocardiography machine] and one new echo[cardiography] machine but they are not usually used in neonates. No one on staff is currently trained in foetal echo.” – Hospital 1*
>
> *“Mongolia is in the process of setting up nationwide pulse oximetry screening*… *However, the programme is struggling to provide the follow-up echocardiogram to screen-failed babies as neither the ultrasound equipment nor suitable trained staff are readily available.” – Meeting with MoH*
>
> *“Abnormalities of the heart are detected by clinical examination and pulse oximetry (since 2018, however, the sensors have been broken for the last few months)*… *The mother is advised to go to the National Maternity and Child Health Centre for further monitoring after discharge from Amgalan.” – Hospital 5*

Staff expressed frustration at long delays caused by broken equipment and lack of repair support:

> *“The main issue facing hospital staff is the lack of maintenance contracts for equipment. When equip. breaks, the hospital is reliant on donation of new technology to replace the broken which can take a significant amount of time.” – Hospital 3*

Lack of digital compatibility between hospital systems further stressed coordination efforts:

> *“One barrier they are facing is the different systems used by different hospitals, and the lack of integration between those systems.” – Hospital 3*

### 4.3 Outer Setting

#### Sub-Domain: Local Conditions

Policy and political leadership aligned with innovation and nationwide screening goals. High-level stakeholders emphasised the importance of early-life interventions and digital infrastructure:

> *“Public health and national screening programmes are now a focus of the presidential office. Health and quality of life are being addressed with multiple screening programmes early in life.” – Presidential Palace Meeting*
>
> *“The Head of [Ministry of Health] MOH Research has been leading the nationwide X-ray screening programme since 2022*… *difficulties*… *needed to support the screening programme*… *turned to AI*… *still hard to connect all hospitals as not all had a [Picture Archiving and Communication System] PACS system or had incompatible eHealth software*…*” – Ministry of Health Meeting*

These factors indicate strong national momentum for implementing new technologies.

### 4.4 The Innovation

Findings mapped to two Innovation sub-domains: Innovation Design and Relative Advantage.

#### Sub-Domain: Innovation Design

A range of feedback on the prototype device design was collected during observations of scanning sessions. Overall, the device was well-received by members of the research team. Areas for improvement were focused on three issues: the angle of the device needed to image the apex of the heart, the size of the device, and the application interface.

> *“The clinicians are happy with the quality of the scan but the probe is requiring a significant amount of manipulation in order to get a ‘good’ or appropriate image.” – Hospital 3*
>
> *“The first baby scanned is a few hours old and is very small. The medical team have trouble positioning the probe in a way that avoids the baby’s head.” – Hospital 3*
>
> *“The scanning application took a very long time to save scans and the [User Interface] felt cumbersome.” – Hospital 1*

There were instances of babies becoming unsettled during scans, however this is not unusual for newborns and young children during exams.

#### Sub-Domain: Innovation Relative Advantage

The device’s compatibility with limited infrastructure and minimal training requirements were viewed as significant advantages:

> *“The Bloom Standard device doesn’t rely on a PACS system as it utilises cloud-based data transfer*… *uses a simplified image screening protocol as it does not need to make an in-depth diagnosis; this is still done by the expert via echocardiogram.” – Meeting with MoH*.

## 5 Discussion

To our knowledge, this study is the first to utilise the CFIR 2.0 to assess the implementation of a novel medical device *during* device development and aims to address a critical gap in published implementation science literature. Whilst theories and frameworks for adopting innovations in healthcare have been developed, these are typically applied after the innovation has been designed and implemented [35].

Rather than attempt to improve implementation after the fact, our research team aim to gather evidence of implementation barriers and facilitators in tandem with device development and design. Our findings demonstrate that early use of CFIR can help identify system-level, organisational, and individual factors that may influence future adoption, leading to improvements in both product design and implementation strategy.

Across four CFIR domains, we identified several favourable conditions for implementation. At the individual level, motivation among clinicians was high, with enthusiasm for the device observed across all sites. Within the inner setting, hospitals showed strong alignment with the innovation’s mission, and many had active collaborations with international partners. Importantly, national leadership and Ministry of Health priorities were also aligned with early screening and digital health, suggesting a supportive outer setting.

However, major barriers remain. The most critical are infrastructural; variability in ultrasound availability, lack of maintenance systems, limited training in neonatal echocardiography, and incompatible eHealth systems. These mirror common structural challenges found in other LMIC implementation studies [25, 26]. Critically, these findings highlight the need for device designs that are robust, minimally dependent on maintenance, and without reliance on hospital-specific digital platforms. This aligns with broader calls for context-responsive medical device innovation in LMICs [18, 20].

Our findings suggest that successful implementation in this context would require not only an intuitive and durable device, but also a deployment model that accounts for human resource limitations. This could include task-shifting to less specialised health workers and cloud-based remote expertise, both of which are approaches increasingly used in addressing global health challenges [36-39].

Whilst some of the identified barriers in this study are context-specific, others – such as the lack of eHealth integration – are widely documented in both LMIC and high-income health systems; a 2021 OECD survey of 27 participating countries identified only 15 that had a unified national eHealth system [40], with the Netherlands and the United States of America among the countries that self-reported as having no mandated national or sub-national eHealth systems in place. As such, the study contributes to the literature by demonstrating how early, structured field observation can help pre-empt implementation barriers in a wider range of environments.

The use of CFIR prospectively, especially with non-intrusive observational methods, also raises important methodological considerations. Our findings show that observational field notes, when thematically analysed and mapped to CFIR domains, can generate useful insights even before formal implementation begins. Observational findings that are subsequently explored and mapped through first-hand experience (e.g., via interviews with those on the implementation front line) could provide an even richer analysis.

Finally, deductive mapping of our inductively-identified themes to CFIR domains improved the clarity of our findings, supporting the identification of individual, organisational, and system-level considerations that may otherwise have been overlooked or under emphasised. By using a pre-defined and well-established framework to structure our results, we facilitate comparison with other CFIR-based studies and wider contribution to the research field.

In summary, this study demonstrates that early implementation assessment of prototype medical devices using CFIR is feasible and informative. The findings highlight the importance of aligning product design with local infrastructure, workforce realities, and systemic limitations from the outset. Further work is needed to operationalise these insights into an actionable implementation strategy.

### Innovation Partner Perspective on Early Implementation Assessment

*The following section reflects the views of the innovation partner and illustrates how early implementation assessment can influence product development and future implementation strategy planning in practice*.

Industry innovators often omit, diminish or delay the identification of critical factors that impact successful device development and implementation, including user-centred design, implementation science, environmental, and human factors. Knowledge limitations around low-resource or global health environments are likely a contributing factor as to why these considerations are not well-incorporated into the problem-definition, ideation and early prototyping stages.

A methodological approach using the CFIR, as outlined here, may assist industry innovators in systematically identifying and applying these critical considerations earlier in the device development pipeline.

The industry team note the following as the highest-impact design and implementation factors identified in this research.

### 1) Individuals

#### Existing Awareness/Design Impacts

Sustained innovation requires committed technology champions and trusted clinical partners. The technology Research & Development team, clinical advisors, and Co-Principal Investigators had prior global health experience and established relationships with key clinical and public health stakeholders in the target setting, informing early design decisions and facilitating engagement.

#### Steps Taken to Support Future Implementation Success

The CFIR-based analysis further highlighted the criticality of developing a strong trust fabric and respectful working clinician partnerships. Being open to routine additions to clinical oversight, collaborations and ongoing user feedback can help mitigate potential for bias and flag unidentified user needs for further development.

### 2) Inner Setting

#### Existing Awareness/Design Impacts

The innovation was intentionally designed for resource-constrained environments using WHO technical specifications and target product profiles, including durability, affordability, and optimized battery performance. While geographic challenges, patient access, and infrastructure barriers were anticipated, additional constraints emerged during field testing. Unanticipated findings are often uncovered during field testing and training, especially for more novel innovations.

#### Steps Taken to Support Future Implementation Success

Iterative field-based data collection was critical to refining hardware and software architecture, particularly to reduce dependence on traditional eHealth infrastructure in remote settings. It is critical for innovation partners to approach this process with an open-mind and a willingness to iterate and mitigate as challenges arise. As identified in the CFIR analysis, this research emphasised the need for software architecture that is designed to mitigate the need for traditional eHealth system infrastructure in the most remote and resource-constrained settings.

### 3) Outer Setting

#### Existing Awareness/Design Impacts

The technology innovation team had a strong, preexisting background in public health, referral planning, training and implementation, focused on LMICs. Pre-existing partnerships with health institutions and key stakeholders in the research setting enabled alignment with national policy priorities, patient care and referral systems throughout the development stages.

#### Steps Taken to Support Future Implementation Success

The analysis highlighted the importance of sustained relationships across Ministries of Health, NGOs, and regional medical societies, while remaining adaptable to evolving policy and infrastructure contexts over time. Moving forward, the industry team will focus on continuing to develop key networking opportunities, whilst having the flexibility to acknowledge and respond to changes in leadership, infrastructure investments or health priorities over time.

### 4) Innovation

#### Existing Awareness/Design Impacts

Multidisciplinary engineering, clinical, regulatory and health economics teams collaborated with in-country partners over three years helped inform iterative hardware and software development. Understanding that eHealth systems and interoperability were challenging in the region, information architecture was intentionally designed to address regional data interoperability gaps, support stakeholder adoption and potentially ease transitions for developing eHealth systems.

#### Steps Taken to Support Future Implementation Success

This was the first clinical site to test a fully automated, mechanical sweep solution for the target patient population. The long-term feedback and iteration cycle referenced above reduced the learning curve among stakeholders and supported the overall reception of the prototype interaction and testing.

Field testing of the fully automated mechanical sweep solution identified opportunities for further optimization in device form factor, sensor scanning angles, and software interface features. Rapid iteration cycles, implemented within 35 days, enabled responsive refinement and retesting.

#### Feedback Summary

The CFIR is increasingly powerful when utilized by innovators, particularly for novel technologies. Prospective application of CFIR throughout the development lifecycle strengthens early identification of implementation risks and design limitations. Flagging areas for improvement, both systemic and/or technology based, can directly impact adjacent CFIR domains. By recognizing the interdependence of CFIR domains, industry teams can better align with academic, clinical, and public health partners, thereby reducing barriers to adoption and accelerating responsible implementation.

## 6 Limitations

Due to the nature of observational field notes, this study is inherently biased towards the perspective and interpretation of the lead researcher. Whilst all efforts were made to write field notes that were representative of the environment and situations witnessed, the opinions and views of others can only be assumed based on their behaviour.

## 7 Conclusions

This study demonstrates the feasibility and practical application of the CFIR for evaluation of implementation factors for a prototype medical device still in development. By applying CFIR prospectively, we were able to identify key barriers and facilitators that may affect future use of the device in a low-resource setting.

This work supports the argument that implementation planning should begin early in the development process, particularly in low-resource environments where infrastructure constraints can significantly impact uptake. By using implementation science methods to inform both product design and strategic planning, there is an opportunity to develop technologies that are better suited to the settings in which they will be used.

The methodology employed in this study offers a practical technique for gathering early implementation insights without the need for large-scale data collection. These findings will inform the next phase of research, which will involve interviews with clinical stakeholders and further exploration of the issues raised in this study.

In the future, the research team aim to address this limitation by conducting interviews with staff and key opinion leaders to explore the areas identified in this study and collect first-hand accounts of their lived experiences.

## 11 Author Contributions

**ER**: Conceptualisation; Methodology; Investigation; Writing – Review & Editing; Supervision; Funding Acquisition

**MM**: Methodology; Formal Analysis; Data Curation; Writing – Original Draft

**UB**: Resources; Writing – Review & Editing; Project Administration

**ND**: Investigation; Writing – Review & Editing

**AS**: Writing – Original Draft; Project Administration

## 12 Data Availability Statement

The data that support the findings of this study (field notes) are not publicly available due to their descriptive nature and institutional sensitivities but may be shared on a case-by-case basis upon reasonable request.

## Footnotes

1 Favourable ethical approval provided by the Research Ethics Review Committee of the Mongolia National University of Medical Sciences

