## Supplementary File 1 for "Designing for Success: A Prospective Evaluation of Implementation Factors Affecting a Prototype Novel Medical Device in a Low-Resource Environment Using the CFIR 2.0" (from another version of this preprint: version 1, 2026-02-03)

### Hospital Key

| Hospital Identifier | Key Contextual Notes |
|---|---|
| Hospital 1 | Remote, general hospital located in the Tov province.<br>Visit conducted in Obstetrics and Gynaecology department. |
| Hospital 2 | Large, specialist maternity hospital located in the capital Ulaanbaatar.<br>General tour of hospital conducted and seminar attended. |
| Hospital 3 | National level hospital for maternal and child health, divided into specialist departments.<br>Located in the capital Ulaanbaatar.<br>Complete tour of facilities conducted. |
| Hospital 4 | Large, but remote hospital located in the Tov province. Not specialised to maternal or infant care.<br>Complete tour of facilities conducted. |
| Hospital 5 | Large, specialist maternity hospital located on the outskirts of Ulaanbaatar.<br>Complete tour of facilities conducted. |

### Note Taking Template

**Date:** **Location:**

**Attendees:**

**Description of activity:**

**Pictures/Videos: [ Y ] [ N ]**

**Raw Notes incl. quotes, conversation summaries etc:**
(be mindful of all 5 senses)

**Overview/Summary:**

**Reflective Thoughts:**

**Questions/Follow-up Items:**
